# Predicting Intensive Care Readmission Among Hospitalized Children

**DOI:** 10.64898/2026.05.15.26353330

**Authors:** Ahmed Arshad, Kyle A. Carey, Latasha A. Daniels, Priti Jani, Emily R. Gilbert, L. Nelson Sanchez-Pinto, Anoop Mayampurath

## Abstract

**Objective:** Readmissions to the PICU are associated with increased morbidity and mortality. A prediction model that can identify children at risk of readmission at the time of transfer can allow providers to intervene and potentially improve patient outcomes. The objective of this study was to derive and validate machine learning models to predict PICU readmission at the time of transfer.

**Design:** Retrospective observational cohort study

**Setting:** Three quaternary care PICUs in the city of Chicago

**Patients:** All children admitted to the PICU between 2012 and 2019.

**Measurements:** The primary outcome was unplanned readmission to the PICU within 48 hours of transfer to the inpatient ward. Predictor variables included vital signs, patient characteristics, and laboratory results. We developed and externally validated four models to predict PICU readmission: logistic regression, elastic net, random forest, and XGBoost.

**Main Results:** This study included 35,601 patients, with readmission rates ranging from 2.2 - 3.7% by site. The performance of models during internal validation was consistent at the three sites, with the area under the receiver operating characteristic (AUC) values between 0.70 and 0.73 and no difference across the four models. Model performance decreased significantly during external validation (AUCs of 0.60 – 0.69). The variables most important to the prediction differed at each site.

**Conclusion:** Machine learning models for predicting readmissions to the PICU have limited generalizability. Locally derived models demonstrated modest performance in our study and could potentially inform provider decision-making if prospectively validated. Externally developed models are unlikely to perform well at predicting PICU readmissions.

## INTRODUCTION

Children who are transferred from the pediatric intensive care unit (PICU) to the acute care ward and subsequently readmitted are at higher risk of morbidity and mortality compared to patients who were not readmitted.^1-4^ PICU readmission rates have been reported to be approximately 4%, with children who are readmitted to the PICU likely to be younger, chronically ill, and have an unscheduled index admission.^1-5^ PICU readmissions are a potentially preventable source of worse patient outcomes, and are often used as quality metric by hospital systems to improve patient safety and health.^6-8^

Pediatric hospitals have limited options to identify children at risk for PICU readmission, such as using early warning scores (usually targeted towards deterioration in the ward)^9^ or adapting adult scoring systems to the pediatric population.^10^ Linton and colleagues developed a data-driven score using retrospective data at their institution, but the score included subjective assessments and thus has limited generalizability.^11^ In adult patients, machine learning (ML) approaches have been shown to be superior to clinical standards in predicting ICU readmission after transfer, ^12-17^ however, a similar approach has yet to be explored in children.

A significant concern regarding ML-based clinical prediction models is their limited generalizability outside of the institutions where they were derived.^18-20^ A scoping review of ML to predict adult and pediatric readmissions demonstrated that half of the included models were derived using data from a single health system and less than 10% were externally validated.^21^ While internal validation is a good indicator of model reproducibility, it is not a guarantee of external generalizability. This was recently illustrated during the external validation of a proprietary sepsis prediction model that had significantly worse performance than in the original derivation cohort.^22^

The aim of this study was to explore the feasibility of using ML-based models to predict unplanned readmissions to the PICU at the time of transfer to the inpatient ward across multiple sites. We hypothesized that ML approaches could predict readmissions earlier and with greater accuracy in internal and external settings than logistic regression.

## MATERIALS AND METHODS

### Setting and Study Population

We conducted an observational cohort study of all children admitted to the PICU at Comer Children’s Hospital at The University of Chicago (UC), Ann & Robert H. Lurie Children’s Hospital (Lurie), and Loyola Medicine Children’s Hospital (Loyola) from January 1, 2012 to December 31, 2019. Patients older than 18 years on admission to the PICU and those who expired during their ICU stay were excluded from this study. Birth encounters for newborn patients were also excluded. The study received approval from local Institutional Review Boards (UC IRB #18-0645; Title: Novel Predictors of Clinical Deterioration in Hospitalized Children; Approved: 05/14/2019. Lurie IRB#2020-3868; Title Characterizing and Predicting Clinical Deterioration at Lurie Children’s Hospital, Approved 07/06/2022. Loyola IRB# 215464; Title: Developing Models to Predict Clinical Deterioration in Children; Approved: 10/17/2022) and was conducted following ethical standards set at the institutions and the Helsinki Declaration. Data were collected from the electronic health record (EHR; Epic, Verona, WI) and hospital administration databases at each institution.

### Outcome Variable

The primary outcome was unplanned PICU readmission, defined as a physical transfer from the PICU to the pediatric ward and back to the PICU within 48 hours during the same hospitalization. The time of transfer was determined using the Admission Discharge Transfer database within the EHR, which was used to calculate the patient location for each EHR observation. Only direct transfers from the pediatric ward to the PICU were considered as readmissions for this study. Patients who returned to the PICU from a procedural service, either from the ward or the PICU, were not considered to have the outcome of interest.

### Predictor Variables

Predictor variables included patient age, vital signs, mental status, laboratory test results, number of prior comorbid conditions (categorized as 0, 1, >1), location of ICU admission (emergency, ICU, ward, or other), fraction of inspired oxygen (FiO2), and critical events during PICU stay (i.e., invasive mechanical ventilation and vasoactive infusions). Our predictor variables were chosen based on measurements of patient physiology taken as part of standard care, the need to minimize collinearity between certain laboratory results, including events in the ICU that are likely to impact future deterioration, and adjusting for patient medical history. Prior comorbidities were derived using the Pediatric Complex Chronic Condition (PCCC) classification system.^23-25^ Briefly, we used a patient’s diagnosis codes from their prior admissions to calculate the number of prior comorbidities according to the PCCC classification criteria. The final value measured closest to the time of transfer from the PICU was used for each predictor variable. Missing values were handled by first carrying forward the last known observations, followed by site-specific median imputation (for internal validation) or medians in the derivation data (for external validation) if no values were recorded. No other processing was conducted across all sites.

### Prediction Modelling

Logistic regression, elastic net, random forest, and gradient-boosted (XGBoost) models were derived and validated at each site. A logistic regression model is a standard model used for binary classification problems. The elastic net model utilizes a logistic regression framework but adds a regularization term to minimize the impact of collinearity between the variables, leading to a more generalizable model. Random forests and XGBoost models are tree-based ensemble machine learning approaches where several decision trees are first created and then the final prediction is made by aggregating the result of individual tree predictions. Therefore, they can use interactions between variables more effectively to drive prediction than regression approaches, while the ensemble approach ensures generalizability. While random forests build uncorrelated decision trees, i.e., they vary in variables and observations used, XGBoost models build sequential trees that progressively minimize training error. Each model comes with a set of hyperparameters that control the learning. In this study, hyperparameter optimization was performed using five-fold cross-validation. The full list of tuned hyperparameters and optimal values chosen for the internal validation models are depicted in **Supplementary Table 1**. For internal validation, patient encounters from each site were split longitudinally into derivation (2012 – 2017) and validation (2018 – 2019) cohorts. Each model was trained on the internal derivation cohort and subsequently tested on the internal validation cohort. Our primary metric for model comparison was the area under the receiver operating characteristic curve (AUC). We used DeLong’s test to conduct statistical evaluation across models. External validation was performed after training a model on the entire cohort of one site and validating at the two other sites.

**Table 1:** Patient characteristics, ICU admission vitals, admission locations, and number of prior comorbidities across three sites.

|  |  | **UC**<br>**(n=11,712)** | **Loyola**<br>**(n=6,350)** | **Lurie**<br>**(n=17,539)** |
| --- | --- | --- | --- | --- |
| **Age, median (IQR), years** |  | 4 (1-6) | 5 (1-6) | 4 (1-11) |
| **Sex, n (%)** | **Female** | 5322 (45.4 %) | 2641 (41.6 %) | 7831 (44.6 %) |
|  | **Male** | 6390 (54.6 %) | 3709 (58.4 %) | 9739 (55.4 %) |
| **Race, n (%)** | **White** | 2934 (25.1 %) | 2955 (46.5 %) | 6936 (39.5 %) |
|  | **Black** | 7251 (61.9 %) | 1621 (25.5 %) | 3797 (21.6 %) |
|  | **Other** | 1527 (13.0 %) | 1774 (27.9 %) | 6840 (38.9 %) |
| **Ethnicity, n (%)** | **Hispanic** | 1382 (11.8 %) | 2591 (40.8 %) | 5875 (33.4 %) |
|  | **Non-Hispanic** | 9915 (84.7 %) | 3641 (57.3%) | 11698 (66.6 %) |
| **Heart Rate (beats/min ± SD)** | | 122 ± 29 | 123 ± 30 | 117 ± 25 |
| **Respiratory Rate (breaths/min ± SD)** | | 29 ± 11 | 30 ± 13 | 27 ± 10 |
| **Systolic BP (mmHg ± SD)** | | 102 ± 20 | 107 ± 17 | 106± 14 |
| **Diastolic BP (mmHg ± SD)** | | 58 ± 17 | 61 ± 16 | 66 ± 14 |
| **Oxygen Saturation (% ± SD)** | | 98 ± 4 | 97 ± 4 | 98 ± 3 |
| **Temperature (F ± SD)** | | 98.1 ± 1.4 | 99.1 ± 1.6 | 98.3 ± 1.6 |
| **Encounter Admission Source, n (%)** | **ER** | 5259 (44.9 %) | 2775 (43.7 %) | 7436 (42.3 %) |
|  | **ICU** | 4615 (39.4 %) | 2848 (44.9 %) | 6115 (34.8 %) |
|  | **Ward** | 573 (4.9 %) | 447 (7.0 %) | 1463 (8.3 %) |
|  | **Other** | 1265 (10.8 %) | 280 (4.4 %) | 2559 (14.6 %) |
| **ICU Admission Source, n (%)** | **ER** | 4258 (36.4 %) | 2229 (35.1 %) | 5233 (29.8 %) |
|  | **ICU** | 4418 (37.7 %) | 2794 (44.0 %) | 5911 (33.6 %) |
|  | **Ward** | 1663 (14.2 %) | 831 (13.1 %) | 3682 (21.0 %) |
|  | **Other** | 1373 (11.7 %) | 496 (7.8 %) | 2747 (15.6 %) |
| **Two or more Prior Comorbid Conditions, n (%)** |  | 3829 (33.0 %) | 1572 (24.8 %) | 13871 (78.9 %) |

The relative contribution of each variable for predicting PICU readmission was compared across the three site-specific models. Variable importance was assessed using the magnitude of coefficients for regression-based models (logistic regression and elastic net) or the relative position across trees for tree-based models (random forest and XGBoost). Analyses were conducted using R version 3.3 (R Project for Statistical Computing) with a two-sided p-value < 0.05 denoting statistical significance.

## RESULTS

### Study Population

Across the three sites, there were a total of 35,601 PICU pediatric admissions who survived till PICU discharge, among which there were a total of 23,408 transfers from PICU to the ward (UC: 8,245 [70%], Loyola: 3,808 [60%] Lurie: 11,355 [65%]). Distributions for demographics and other patient characteristics from each site are presented in **Table 1**. The three sites had similar distributions of age and sex. There was a significantly larger proportion of black patients and a smaller proportion of Hispanic patients at UC compared to the other two sites. The percentage of patients with two or more prior comorbid conditions at Lurie was twice as high as the other sites. Initial vital signs for patients across the three sites were similar. UC had 435 readmissions (3.7 %), Loyola had 146 (2.3 %), and Lurie had 389 (2.2 %) during the study period.

### Model Performance

**Table 2** presents the results of the internal validation of prediction models at each site. The logistic regression model at UC demonstrated limited performance in predicting PICU readmissions (AUC 0.64, 95% CI: 0.56-0.71). The odds ratios and 95% CI of the logistic regression model are given in **Supplementary Table 2**. Adding PICU length of stay did not improve the logistic regression model’s performance at UC (AUC 0.63 vs. 0.64, P=0.82). The XGBoost model performed the best at UC, although the performance difference was not statistically significant when compared to the logistic regression model (XGBoost AUC 0.70, 95% CI: 0.63-0.77 vs. logistic regression AUC 0.64, 95% CI: 0.56-0.71, P=0.17). At the optimal cutpoint on the receiver operating characteristic curve, as determined by Youden’s index, the XGBoost model had a sensitivity of 60%, a specificity of 73%, and a number needed to alert 25 in the UC internal validation dataset. The best-performing model at Loyola was the random forest model, which again was not statistically significant from the performance of the logistic regression model (random forest AUC 0.73, 95% CI: 0.63-0.82 vs. logistic regression AUC 0.65, 95% CI: 0.47-0.84, P=0.52). At Lurie, the logistic regression model had the best performance (AUC 0.70, 95% CI: 0.66-0.74). The areas under the precision-recall curve for the best-performing internal validation model at each site are - UC (XGBoost): 0.05, Loyola (random forest): 0.01, and Lurie (logistic regression): 0.04. The best external validation performance for each site (AUC range 0.60 – 0.67) was lower than internal validation (AUC range 0.70 – 0.73) (see **Figure 1)**. The performance of the best-performing external model at UC, i.e., the Lurie XGB model with an overall AUC of 0.67, was similar to or better than model performance when we considered subpopulations based on the number of prior comorbidities (0 prior comorbidities AUC: 0.67; 1 prior morbidity AUC 0.62; >1 prior comorbidity AUC: 0.67). Detailed results for each model that was externally validated are presented in **Supplementary Table 3**. The sensitivity, specificity, positive predictive value, and F1 scores for the best-performing internal and external validation models at each are given in **Supplementary Tables 4 and 5**, respectively.

**Table 2:**
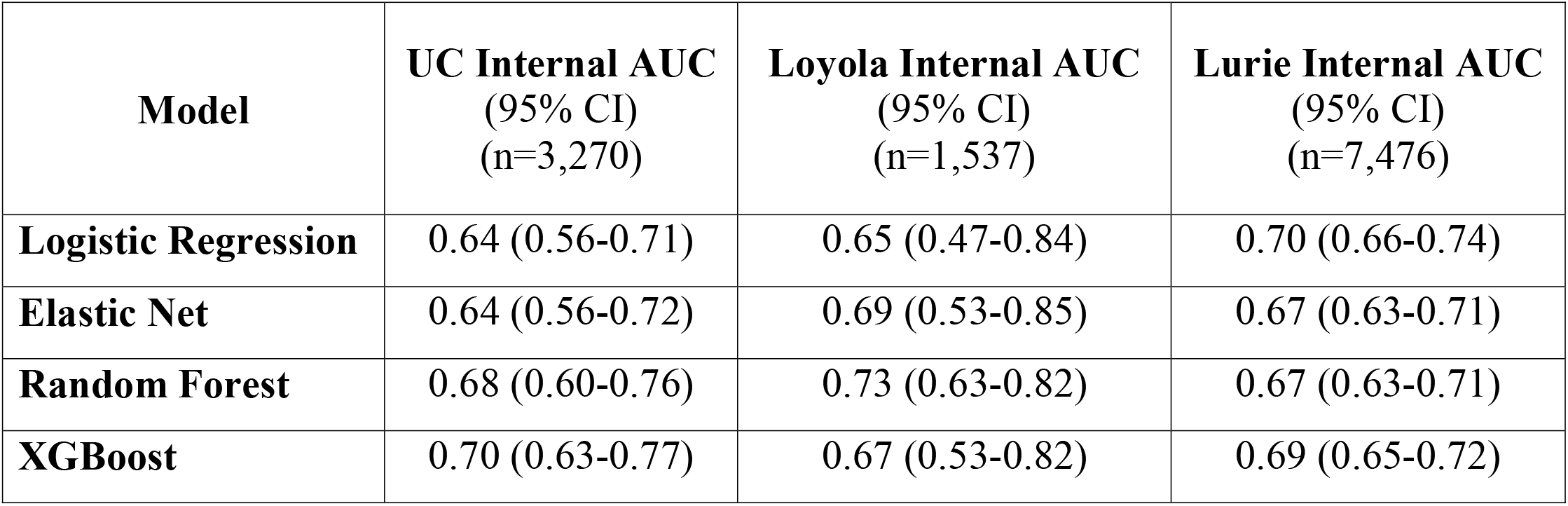
Internal validation AUCs.

**Figure 1.**
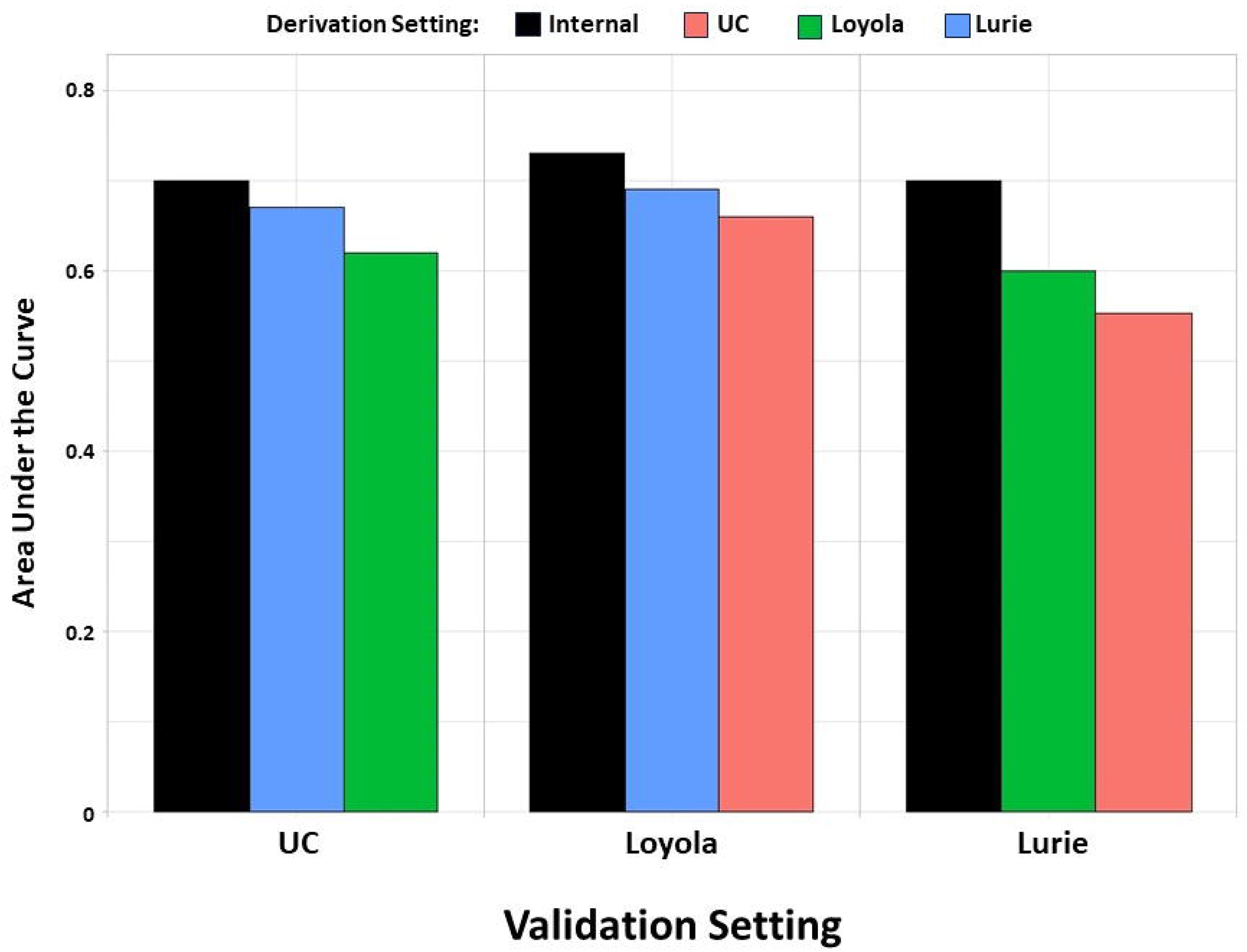
Comparison of internal models with the best performing external model at each site

Analysis of variable importance for the best-performing models at each site revealed differences in the importance of predictors (see **Figure 2**). Variables important for the best-performing XGBoost model from UC were vital signs followed by lab results. At Loyola, interventions and patient factors, including critical events, mental status, comorbidities, and admission source, were the most important predictors for the random forest model. Finally, the logistic regression model trained at Lurie had a mix of patient factors, vital signs, and laboratory results as the most important predictors.

**Figure 2.**
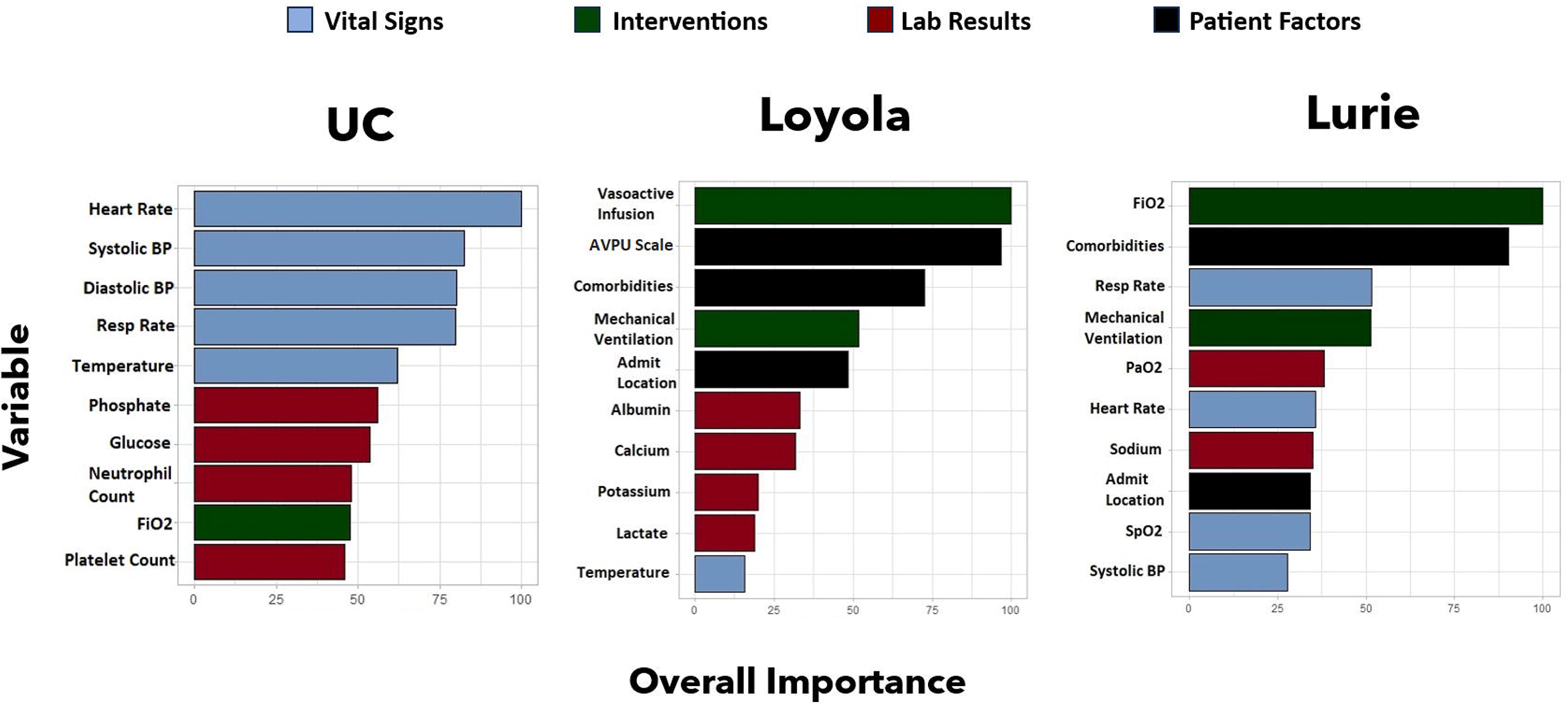
Relative importance of variables for the best performing prediction model at each site

## DISCUSSION

In this study, we describe the performance of machine learning models to predict unplanned readmission to the PICU within 48 hours of transfer in both internal and external settings. Our models predicted unplanned readmission to the PICU with limited performance during internal validation, with machine learning models demonstrating no clear superiority over logistic regression models. To our knowledge, no similar model exists for pediatric patients, however, performance was on par with models for adult patients.^14,16,26^ Further, we observed that model performance worsened during external validation. Our results indicate that while there is a possibility that real-time data can stratify the risk of readmission for each patient prior to transfer from the PICU, PICU readmission is a problem dependent on local processes and cannot be generalized easily.

Readmissions in the pediatric critical care population are difficult to predict in part due to their low frequency. However, for each patient, readmission results in significantly worse outcomes.^1-5^ The ability to accurately predict the risk of readmission prior to transfer creates the opportunity for intervention to mitigate potential adverse outcomes. Within our study, we noted few instances of significant increase in performance between logistic regression and more advanced ML models. This implies that for the purpose of predicting PICU readmission, more complex ML modelling is not superior to logistic regression. Currently, there are no ML models that predict readmission to the PICU to compare our results with. The Prediction of PICU Early Readmissions (PROPER) score adapted an adult score to predict PICU readmissions with an AUC of 0.795.^10^ While the AUC for the PROPER score is higher than reported for our models, there are a few important distinctions to consider. First, due to the case-control design of the PROPER study, the system was tested on a much smaller and controlled sample than the ones our models were validated on. Second, the PROPER score does not include laboratory results compared to the more comprehensive list of variables included in our models. Third, the PROPER score includes functional scoring systems that are manually calculated on admission which may have decreased utility at the bedside compared to a prediction model that provides a real-time risk of readmission score at the time of transfer. Finally, most of the factors included in the PROPER score are non-modifiable, and therefore provide limited opportunities for intervention. In contrast, the variable importance results for our models highlight specific vital signs and lab results that influence the prediction and provide actionable information to clinicians.

This is the first study to apply ML to predict readmissions in a pediatric population and report external validation across three different sites. Our results show that extending from logistic regression to machine learning models did not improve performance in any of the three sites. It is possible that the outcome of PICU readmission may not be dependent on non-linear interactions among variables. However, the limited number of events or exclusion of potential confounders could also play an important role. Our results also show that external validation across the three sites in this study revealed lower performance when compared with internal validation. Notably, lower external validation performance was observed across all site-specific models, regardless of the cohort used for derivation or the machine learning approach. The drop in performance could have multiple potential explanations. First, this may be due to differences in the patient cohorts at each of our three sites, especially with regard to prior comorbidities. It is possible that PICU readmission may be sensitive to different diseases and treatment options offered at each site. Second, the criteria for PICU admission could be unique to each site. While there are conditions that must always be treated in a critical care setting, hospital policy and resources may govern the provision of interventions in the wards.

Our study suggests that attempting to implement externally derived models without local testing could lead to significant inaccuracies, thereby indicating the need to balance the ability of a hospital to train a model on site-specific data against the limited performance of an externally trained model. Training an in-house model requires processes and expertise in EHR data extraction, cleaning, and model building. While most regression-based external models can be implemented easily, implementing a more advanced external model requires technical capabilities. Therefore, it may be in the best long-term interest of hospitals to train PICU readmission models in terms of improved performance and the ability to replicate the workflow for other clinical prediction models. However, it is still possible to localize an externally derived model to ensure adequate performance. For example, De Hond et al. initially reported poor performance during external validation of an ML model that predicted ICU readmission or death within seven days of transfer in adult patients.^27^ However, retraining the model using data from the test site and improved performance to be comparable to the original internal validation. Whether our PICU readmission models could benefit from such retraining at each site is an area of future work. While a single generalizable model capable of good performance and use across multiple institutions would be ideal, different populations and practices could prove challenging for PICU readmissions.^18^ Additionally, a local model might be more actionable and lead to better patient outcomes, when compared with one that compromises clinical utility for generalizability.^28^ PICU readmission model outputs at the time of PICU discharge can be converted to a numeric score indicating the likelihood of being readmitted to the PICU, using approaches similar to prior work,^29^ thereby enabling decision-making at the patient’s bedside. However, operationalizing machine learning models without adequate validation and localization can lead to unnecessary notifications and alert fatigue for providers and create the possibility of patient harm.^22^

Our variable importance analysis revealed different factors that predict readmission across the three sites. The differences in variable importance among the site-specific models could explain the lack of generalizability observed for our models. For example, at Lurie, oxygen-related variables and prior history of comorbidities are the most important variables for predicting readmissions. Factors related to the current severity of illness are most important for predicting readmissions in Loyola, while at UC, the last recorded vital sign readings appear to be the main drivers of predicting readmissions. These differences may be driven by underlying variations in population and practice. It is also important to recognize that some risk factors for readmission are modifiable, while others, such as prior comorbidities, are not. Our results indicate that clinical decision-making to reduce unplanned readmissions should be site-specific. By understanding local factors that are driving the prediction for readmission, providers can decide which interventions will be most significant to mitigating that risk for their patient population.

There are several limitations to our study. While we adopted a multi-site approach to maximize generalizability, our focus on retrospective data limits the evaluation of our results. We hope this study will lead to additional research in this area, including a comparison with the PROPER model in a prospective setting or testing performance in more targeted populations. Validating our findings using large-scale multicenter data, such as from the Virtual Pediatric Systems (VPS), is an important area of future work. Some of our limitations are inherent to the nature of readmissions as an outcome. The overall rate of PICU readmission in our cohort was low. While this is representative of published data for PICU readmissions,^1-5^ it does limit our ability to derive a more accurate statistical model. For example, it is possible that a model trained on multicenter data and then adapted to a local site using transfer learning may perform better than an internally derived model. Another limitation is that our study is retrospective in nature and may be missing other important predictors, such as granular details regarding respiratory support. While we considered the last recorded laboratory values as features in our model, labs taken closer, e.g., 48 hours prior to the time of PICU discharge, may be more predictive of being readmitted to the PICU. Additionally, hospital-level variables that impact readmission were not included in our data. In particular, factors related to patient care outside of the PICU, such as being on high-flow nasal cannula in the ward or in the emergency room, could also contribute to the lack of generalizability. Determining the importance of these variables remains an area of future research.

## CONCLUSION

In conclusion, we developed and externally validated multiple models using vital signs, laboratory results, and patient factors to predict unplanned readmission to the PICU at the time of transfer. Our results make a strong argument for local validation and likely retraining models prior to prospective validation in a clinical setting for detecting patients at risk for PICU readmission.

## Data Availability

All data produced in the present work are contained in the manuscript and supplement

## Acknowledgements

We thank the Center for Research Informatics at the University of Chicago Center and the Informatics and Clinical Research team at Loyola University Medical Center for their assistance in data procurement and cleaning.

Preliminary results for this study were presented and the Pediatric Data and Analytics subgroup of PALISI, we thank them for their helpful feedback.

