## Supplemental Tables 1-5 for "Predicting Intensive Care Readmission Among Hospitalized Children"

**Supplementary Table 1:** List of hyperparameters optimized for each model along with final values chosen for models trained using the internal derivation dataset at UC.

| **Model** | **Parameters** | **Search Values** | **Best value** |
| --- | --- | --- | --- |
| Elastic Net | Lambda | 0, 0.0001, 0.0003, 0.001, 0.005, 0.01, 0.1, 0.5, 1 | 0.1 |
|  | Alpha | 0.00, 0.01, 0.02, 0.3, 0.5, 0.7,1 | 0 |
| Random Forests | Number of trees | 500, 1000, 2000, 2500 | 2000 |
|  | Number of variables available for splitting at each tree node. | 4, 5, 6, 7, 8 | 6 |
| Gradient Boosted Trees | Number of trees | 300, 500, 1000, 1500 | 500 |
|  | Maximum depth of trees | 5, 10, 15, 20 | 15 |
|  | Learning rate | 0.001, 0.01, 0.05, 0.1 | 0.01 |
|  | Fraction of samples supplied to tree | 0.5, 0.7, 1 | 0.7 |
|  | Gamma | 0 | 0 |
|  | Fraction of features supplied to tree | 1 | 1 |
|  | Minimum sum of instance weight | 1 | 1 |

**Supplementary Table 2**: Odds ratios (with 95% CI) for variables used in the logistic regression model derived using internal UC derivation (years 2012-2017) dataset.

| **Variable** | **OR (95% CI)** |
| --- | --- |
| Age | 0.98 (0.94-1.04) |
| Heart Rate | 1.00 (0.99-1.01) |
| Respiratory Rate | 1.03 (1.00-1.06)* |
| Systolic Blood Pressure | 1.00 (0.99-1.02) |
| Diastolic Blood Pressure | 0.99 (0.99-1.02) |
| Oxygen Saturation | 1.04 (0.98-1.14) |
| Temperature | 1.26 (1.06-1.48)* |
| FiO2 | 1.01 (1.00-1.01)* |
| AVPU Scale, ref 0 |  |
| 1 | 0.92 (0.05-4.32) |
| 2 | 6.42 (1.47-1.94)* |
| 3 | - |
| Albumin | 0.62 (0.36-1.08) |
| Anion Gap | 0.99 (0.91-1.07) |
| Bicarbonate | 1.05 (0.99-1.12) |
| Blood Urea Nitrogen | 1.01 (0.98-1.04) |
| Calcium | 0.94 (0.66-1.36) |
| Creatinine | 0.97 (0.53-1.39) |
| Glucose | 1.00 (0.99-1.00) |
| Hemoglobin | 1.00 (0.87-1.15) |
| Lactate | 1.14 (0.83-1.43) |
| MCV | 0.98 (0.95-1.02) |
| Neutrophil | 0.99 (0.98-1.01) |
| PaO2 | 1.00 (0.99-1.00) |
| PCO2 | 0.97 (0.94-0.99)* |
| pH | 0.86 (0.94-1.00) |
| Phosphate | 0.68 (0.53-0.87)* |
| Platelet | 1.00 (1.00-1.00)* |
| Potassium | 1.25 (0.89-1.72) |
| Sodium | 0.98 (0.92-1.04) |
| Troponin | - |
| WBC | 1.00 (0.97-1.01) |
| Prior comorbidities, ref 0 |  |
| 1 | 1.64 (0.68-2.43) |
| > 1 | 1.94 (0.126-2.93)* |
| Encounter admit location, ref ED |  |
| Ward | 1.28 (0.55-2.89) |
| ICU | 0.89 (0.14-3.34) |
| Other | 0.88 (0.25-2.98) |
| ICU admit location, ref ED |  |
| Ward | 1.47 (0.75-2.72) |
| ICU | 1.80 (0.45-1.22) |
| Other | 0.66 (0.17-2.19) |
| Invasive Mechanical Ventilation | 0.59 (0.34-0.98)* |
| Administration of vasoactive medication | 1.11 (0.43-2.48) |

*P<0.05

- indicates ORs could not be determined due to low counts.

**Supplementary Table 3**: External validation of predictive models across the three sites. * denotes the best-performing external model for that testing site across all external validation.

| **Logistic Regression** | | **Testing Site** | | |
| --- | --- | --- | --- | --- |
|  |  | **UC** | **Loyola** | **Lurie** |
| **Training Site** | **UC** | - | 0.58 (0.50 – 0.65) | 0.54 (0.50 – 0.55) |
|  | **Loyola** | 0.54 (0.50 – 0.58) | - | 0.59 (0.56 – 0.62) |
|  | **Lurie** | 0.55 (0.51 – 0.60) | 0.56 (0.49 – 0.63) | - |

| **Elastic Net** | | **Testing Site** | | |
| --- | --- | --- | --- | --- |
|  |  | **UC** | **Loyola** | **Lurie** |
| **Training Site** | **UC** | - | 0.63 (0.56 – 0.70) | 0.55 (0.52 – 0.58) |
|  | **Loyola** | 0.59 (0.55 – 0.63) | - | **0.60 (0.58 – 0.63)*** |
|  | **Lurie** | 0.61 (0.56 – 0.65) | 0.57 (0.50 – 0.64) | - |

| **Random Forest** | | **Testing Site** | | |
| --- | --- | --- | --- | --- |
|  |  | **UC** | **Loyola** | **Lurie** |
| **Training Site** | **UC** | - | 0.64 (0.59 – 0.70) | 0.52 (0.50 – 0.55) |
|  | **Loyola** | 0.60 (0.56 – 0.64) | - | 0.51 (0.48 – 0.54) |
|  | **Lurie** | 0.64 (0.60 – 0.68) | 0.64 (0.58 – 0.70) | - |

| **XG Boost** | | **Testing Site** | | |
| --- | --- | --- | --- | --- |
|  |  | **UC** | **Loyola** | **Lurie** |
| **Training Site** | **UC** | - | **0.66 (0.60 – 0.72)*** | 0.52 (0.50 – 0.55) |
|  | **Loyola** | 0.62 (0.58 – 0.66) | - | 0.49 (0.46 – 0.52) |
|  | **Lurie** | **0.67 (0.63 – 0.70)*** | 0.63 (0.57 – 0.70) | - |

**Supplementary Table 4**: Performance metrics of the best-performing internal model at each site, calculated at the Youden’s Index.

| **Site** | **Model** | **Sensitivity** | **Specificity** | **PPV** | **F1-score** |
| --- | --- | --- | --- | --- | --- |
| UChicago | Gradient-boosted | 60% | 73% | 4% | 0.08 |
| Loyola | Random Forest | 91% | 55% | 1% | 0.02 |
| Lurie | Logistic Regression | 66% | 69% | 4% | 0.08 |

**Supplementary Table 5**: Performance metrics of the best-performing external model at each site, calculated at the Youden’s Index.

| **Testing Site** | **Training Site** | **Model** | **Sensitivity** | **Specificity** | **PPV** | **F1-score** |
| --- | --- | --- | --- | --- | --- | --- |
| UChicago | Lurie | Gradient Boosted | 68% | 59% | 3% | 0.06 |
| Loyola | Uchicago | Gradient Boosted | 69% | 57% | 2% | 0.04 |
| Lurie | Loyola | Elastic Net | 48% | 65% | 3% | 0.06 |
